# The Effect of Occupational Integration on Musculoskeletal Injury in Female Marines in the Fleet: An Epidemiological Cohort Study

**DOI:** 10.64898/2026.02.19.26346637

**Authors:** John J. Fraser, James M. Zouris, Johanna M. Hoch, Pinata H. Sessoms, Andrew J. MacGregor, Matthew C. Hoch

**Affiliations:** Operational Readiness Directorate, Naval Health Research Center, 140 Sylvester Road, San Diego, CA 92106-3521, USA; University of Kentucky Sports Medicine Research Institute, 720 Sports Center Drive, Lexington, KY 40506-0277, USA; Uniformed Services University of the Health Sciences, School of Medicine, Department of Physical Medicine and Rehabilitation, 4301 Jones Bridge Road, Bethesda, MD 20814, USA

**Keywords:** Occupational Injuries, Military Personnel, Sports Medicine, Women’s Health, Public Health

## Abstract

**Introduction:** Musculoskeletal injuries (MSKIs) are ubiquitous in the U.S. military, especially among high-performing service members such as Marines. Given that female service members only started to be assigned to ground combat roles since December 2015, evaluation of sex on MSKI risk in ground combat occupations has not been possible until there was an ample population to study. The purpose of this population-level epidemiological study was to assess (1) if female sex was a salient risk factor for MSKI in Marines serving in different military occupations, including combat arms, and (2) the effects of integration period on MSKI risk among female Marines.

**Materials and Methods:** A population-based epidemiological retrospective cohort study of all U.S. Marines was performed assessing female sex, occupation, and integration period on the prevalence of MSKI from 2011 through 2020. The Military Health System Data Repository was utilized to identify initial healthcare encounters for diagnosed ankle-foot, knee, lumbopelvic-hip, thoracocostal, cervicothoracic, shoulder, elbow, or wrist-hand complex injuries. Prevalence was calculated for female and male Marines in each occupational category (combat, combat support, aviators, aviation support, services) during the pre-integration (2011–2015) and post-integration (2016–2020) periods.

**Results:** During the pre-integration period, 520/1,000 female Marines (*n*=13,985) and 299/1,000 male Marines (*n*=142,158) incurred MSKIs. In the post-integration period, the prevalence increased to 565/1,000 female Marines (*n*=17,608) and 348/1,000 male Marines (*n*=161,429). In the multivariable evaluation of sex, occupation, integration period, and the interaction of sex and occupation on combined MSKIs, only female sex was a significant factor for injury (prevalence ratio [PR]=1.99), with service in ground combat and aviation occupations identified as protective factors when compared with services occupations (PR=0.69). When these same factors were evaluated for specific MSKI outcomes, female sex remained a robust factor in all lower quarter (PR=1.75–2.63) and upper quarter (PR=1.38–2.36) injuries except for shoulder injuries. Service in ground combat and aviation occupations was protective for all lower quarter injuries (PR=0.46–0.71). In the upper quarter, ground combat was protective for all injuries except for elbow injuries (PR=0.67–0.77). Serving as an aviator was a risk factor for cervicothoracic (PR=1.57) and thoracocostal (PR=1.22) injuries and a protective factor for shoulder (PR = 0.73) and wrist-hand (PR = 0.46) injuries. Adjusted risk for lumbopelvic-hip (PR=1.13), ankle-foot (PR=1.53), cervicothoracic (PR=1.19), thoracocostal (PR=1.14), and elbow (PR=1.48) injuries significantly increased during the post-integration period. There was a significant sex-by-period interaction for shoulder injuries alone, with female sex in the post-integration epoch found to be salient (PR=1.26).

**Conclusions:** Female sex was a salient factor for MSKI, with service in ground combat and aviation occupations identified as protective factors when compared with services occupations. In the evaluation of specific MSKIs, female sex remained a robust and significant factor in all lower quarter injuries and upper quarter injuries except for shoulder injuries. There was only a significant sex-by-period interaction for shoulder conditions, with an increased risk of these injuries in female Marines in the post-integration period.

## INTRODUCTION

Musculoskeletal injuries (MSKIs) are highly prevalent in the U.S. military, especially among high-performing service members such as Marines. MSKI episodes occurred at a rate of 1,004.73 episodes per 1,000 person-years in U.S. Marines between December 2016 and August 2021.^1^ In garrison, these conditions pose a substantial threat to operational readiness, affecting service members’ ability to train and deploy to meet unit and national objectives.^1^ MSKIs, which include both macrotraumatic (categorized as injury/poisoning) and repetitive microtraumatic (categorized as musculoskeletal disease) diagnostic categories, are the leading medical conditions in the Military Health System (MHS) and affect more service members than any other category.^2^ This does not change when service members deploy.^3^ When macrotraumatic and repetitive microtraumatic diagnostic categories are combined, MSKIs are the leading medical conditions that result in medical evacuation of deployed service members out of the operational theater.^4^ The most recent finding of MSKI as a leading nonbattle injury and reason for medical evacuation is consistent with the findings reported by Cohen and colleagues in 2010,^5^ who also found that U.S. Marines had the lowest return-to-duty rates following evacuation among the service branches deployed to the Iraq and Afghanistan conflicts.

Female service members have greater injury risk than their male counterparts in the same occupation for various musculoskeletal conditions in the lower extremity.^1,6–11^ This is likely a result of not only biomechanical and physiological factors, but also psychosocial determinants of care-seeking following the onset of symptoms or injury-related impairment.^1,6–14^ The evidence suggests that military occupations not only have varying physical loads and environmental exposures that may affect injury risk, but also unique cultures that may influence care-seeking.^7,9–14^ This is especially true within combat arms occupations in the Fleet Marine Force.

To project power in austere, uncertain, and arduous conditions under the threat of harm at “the tip of the spear,” ground combat occupations are composed of service members that are proactive, mission centered, highly resilient, and have substantial grit. Not dissimilar to care-seeking behaviors in athletes, service cultures within combat arms likely promote self-management and reduce care-seeking following injury.^12^ The cultural factors in ground combat occupations that preclude care-seeking may plausibly counteract the positive care-seeking behaviors typically observed in women. Given that female service members have only been assigned to ground combat roles since December 2015,^15^ evaluation of sex and MSKI risk in ground combat occupations has not been possible until there was an ample population to study. Therefore, the purpose of this population-level epidemiological study was to assess (1) if female sex was a salient risk factor for MSKI in Marines serving in different military occupations, including combat arms, and (2) the effects of the integration period on MSKI risk among female Marines. We hypothesized that female service members would demonstrate greater musculoskeletal injury risk compared with male service members, particularly in combat occupations. Due to the changes in occupational and training exposure following the policy change, which likely had service-wide effects, we anticipated that there would be higher risk of MSKI in female Marines in the post-integration period compared with the pre-integration period.

## METHODS

A population-based epidemiological retrospective cohort study of all service members in the U.S. Marine Corps from 2011 through 2020 was performed assessing female sex, occupation, and integration period on the prevalence of MSKIs. The Military Health System Data Repository (MDR) was utilized to identify relevant healthcare encounters. This study was approved by the Naval Health Research Center Institutional Review Board. The Strengthening the Reporting of Observational Studies in Epidemiology (STROBE) guidelines were used to guide reporting.^16^

Diagnosis codes for MSKI were derived and categorized by regional complex using the Neuromusculoskeletal Epidemiological Outcome (NEO) Matrix.^17^ The database was queried for the number of distinct Marines seen in the MHS on their initial encounter and diagnosed with an ankle-foot, knee, lumbopelvic-hip, thoracocostal, cervicothoracic, shoulder, elbow, or wrist-hand complex injuries.^17^ Prevalence was calculated for female and male Marines in each occupational category (combat, combat support, aviators, aviation support, services) during the pre-integration (2011–2015) and post integration (2016–2020) periods.

Prevalence ratio (PR) point estimates and 95% confidence intervals (CIs), risk difference, attributable risk, and chi-squared statistics were calculated in the assessments of sex and integration period (in female Marines) using Microsoft Excel for Mac 2016 (Microsoft Corp., Redmond, WA) and a custom epidemiological spreadsheet.^18^ In the unadjusted assessment of MSKI risk stratified by body region, male Marines and the 5-year pre-integration period served as reference groups.

Adjusted multivariable models were employed to assess the effects of sex, occupation, integration period, and sex-by-period interaction on counts of musculoskeletal injury for each body region, normalized to the at-risk population. Due to overdispersion of the data, a negative binomial model was employed over a Poisson model.^19^ The regression analyses were performed using the ‘MASS’ package (version 7.3-60.0.1) on R (version 4.3.3, The R Foundation for Statistical Computing, Vienna, Austria). The level of significance was *p* <.05 for all analyses. PR point estimates were considered statistically significant if CIs did not cross the 1.00 threshold. Convergence of *p*-values, effect sizes, and 95% CIs were considered when evaluating significant findings.

## RESULTS

During the pre-integration period (2011–2015), 520 per 1,000 female Marines (*n* = 13 985) and 299 per 1,000 male Marines (*n* = 142,158) incurred MSKIs. In the post-integration period (2016–2020), the prevalence increased to 565 per 1,000 female Marines (*n* = 17,608) and 348 per 1,000 male Marines (*n* = 161,429). The pre-integration and post-integration prevalence in male and female Marines in each occupational category are detailed in Table 1. Supplemental Figure 1 illustrates the changes in MSKI prevalence over time in male and female Marines serving in each occupational category and for all occupations combined.

**Table 1.** Prevalence of Musculoskeletal Injuries (per 1,000 Marines) in U.S. Marines During the Pre-Integration (2011–2015) and Post-Integration Periods (2016–2020), Stratified by Military Occupational Category

| Body region | Sex | Military occupational category |  |  |  |  |  |  |  |  |  |
| --- | --- | --- | --- | --- | --- | --- | --- | --- | --- | --- | --- |
|  |  | Combat |  | Combat support |  | Aviators |  | Aviation support |  | Services |  |
|  |  | Pre-integration | Post-integration | Pre-integration | Post-integration | Pre-integration | Post-integration | Pre-integration | Post-integration | Pre-integration | Post-integration |
| Ankle-Foot | Female | N/A | 171.1 | 150.9 | 204.6 | 62.6 | 208.8 | 131.5 | 186.9 | 173.5 | 214.9 |
|  | Male | 57.9 | 89.0 | 98.2 | 124.5 | 24.0 | 70.3 | 82.7 | 105.5 | 107.2 | 144.7 |
| Knee | Female | NA | 86.7 | 150.8 | 149.5 | 99.9 | 79.8 | 131.4 | 139.4 | 161.5 | 151.1 |
|  | Male | 50.6 | 65.6 | 96.7 | 98.8 | 32.4 | 40.8 | 81.0 | 89.2 | 97.3 | 108.2 |
| Lumbopelvic-Hip | Female | NA | 261.3 | 231.1 | 289.6 | 107.5 | 149.3 | 224.4 | 280.3 | 241.1 | 292.5 |
|  | Male | 48.7 | 69.4 | 85.0 | 109.5 | 63.2 | 64.7 | 163.9 | 98.7 | 85.8 | 111.7 |
| Thoracocostal | Female | NA | 34.1 | 51.6 | 60.0 | 43.5 | 41.6 | 55.0 | 56.9 | 48.9 | 61.6 |
|  | Male | 14.5 | 19.3 | 24.4 | 30.8 | 33.8 | 26.8 | 24.1 | 25.6 | 21.9 | 30.9 |
| Cervicothoracic | Female | NA | 38.2 | 35.6 | 38.5 | 22.2 | 66.9 | 35.3 | 44.5 | 36.7 | 47.2 |
|  | Male | 11.0 | 13.0 | 16.1 | 20.9 | 28.7 | 25.9 | 17.8 | 20.3 | 16.0 | 21.8 |
| Shoulder | Female | NA | 48.2 | 51.5 | 56.1 | 11.8 | 60.9 | 37.1 | 50.0 | 45.0 | 56.9 |
|  | Male | 31.8 | 38.9 | 52.3 | 57.6 | 25.4 | 26.6 | 54.1 | 57.2 | 50.2 | 60.1 |
| Elbow | Female | NA | 33.6 | 8.7 | 13.7 | 15.4 | 6.7 | 8.5 | 15.7 | 10.9 | 14.4 |
|  | Male | 6.9 | 10.6 | 8.6 | 15.0 | 5.6 | 7.5 | 11.0 | 15.0 | 10.1 | 14.9 |
| Wrist-Hand | Female | NA | 41.4 | 43.0 | 49.0 | 26.5 | 13.6 | 66.7 | 63.5 | 50.3 | 66.2 |
|  | Male | 26.6 | 34.8 | 39.9 | 42.7 | 15.8 | 16.2 | 51.6 | 59.7 | 49.3 | 59.0 |

N/A: Not applicable because female Marines were excluded from combat occupations prior to integration.

In the unadjusted assessment of sex on MSKI (stratified by body region) during the post-integration period, female sex was a significant factor for all lower quarter injuries (PR = 1.62–2.93) and thoracocostal, cervicothoracic, and wrist-hand injuries in the upper quarter (PR = 1.26–2.25) (Table 2). In the evaluation of the effects of integration in female Marines, there was a significantly higher prevalence of ankle-foot (PR = 1.29) and lumbopelvic-hip injuries (PR = 1.23) in the lower quarter and all upper quarter injuries (PR = 1.17–1.46) following integration (Table 3).

**Table 2.**
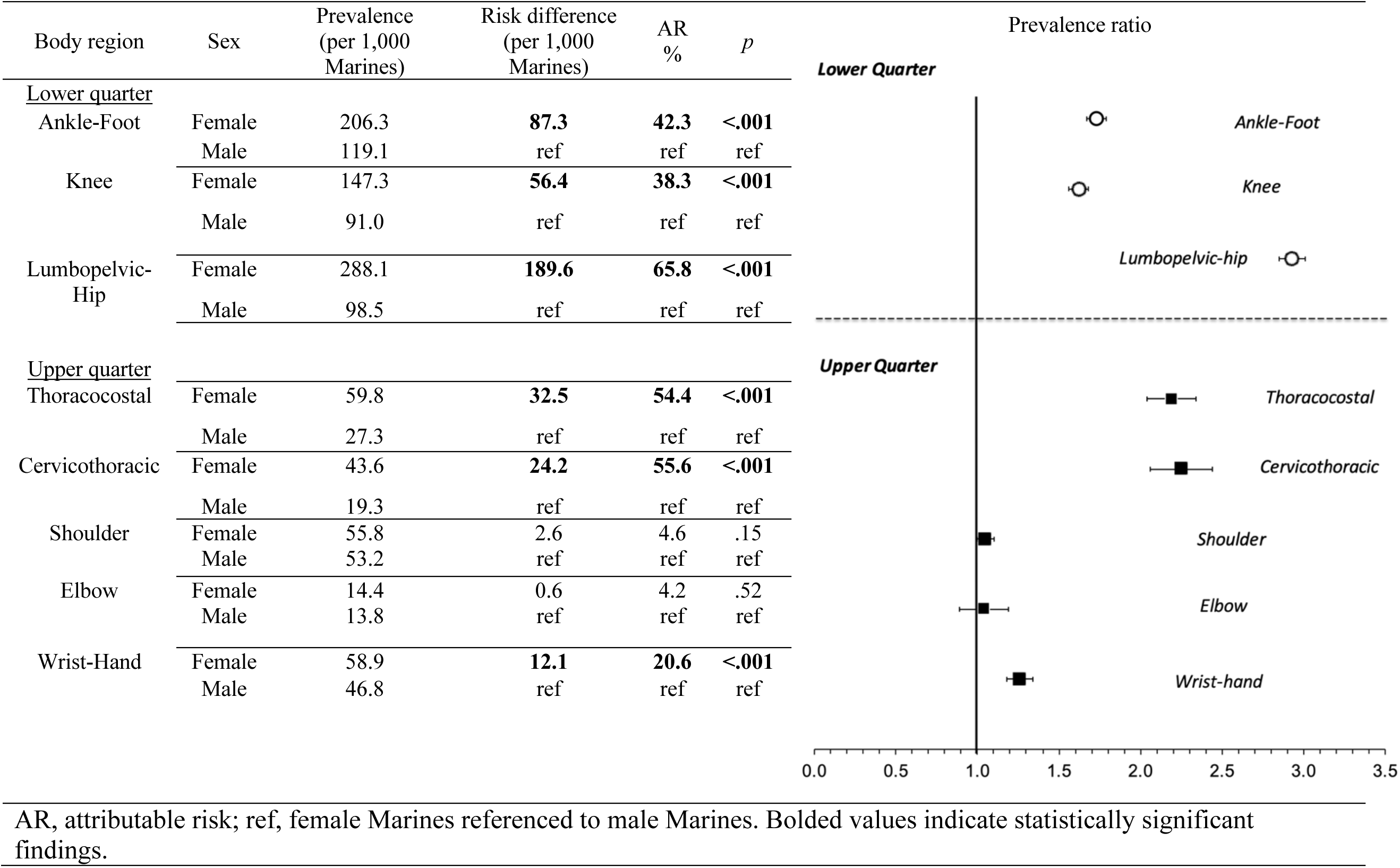
Assessment of Female Sex on Musculoskeletal Injuries, Stratified by Body Region, in U.S. Marines During the Post-Integration Period, 2016–2020

**Table 3.**
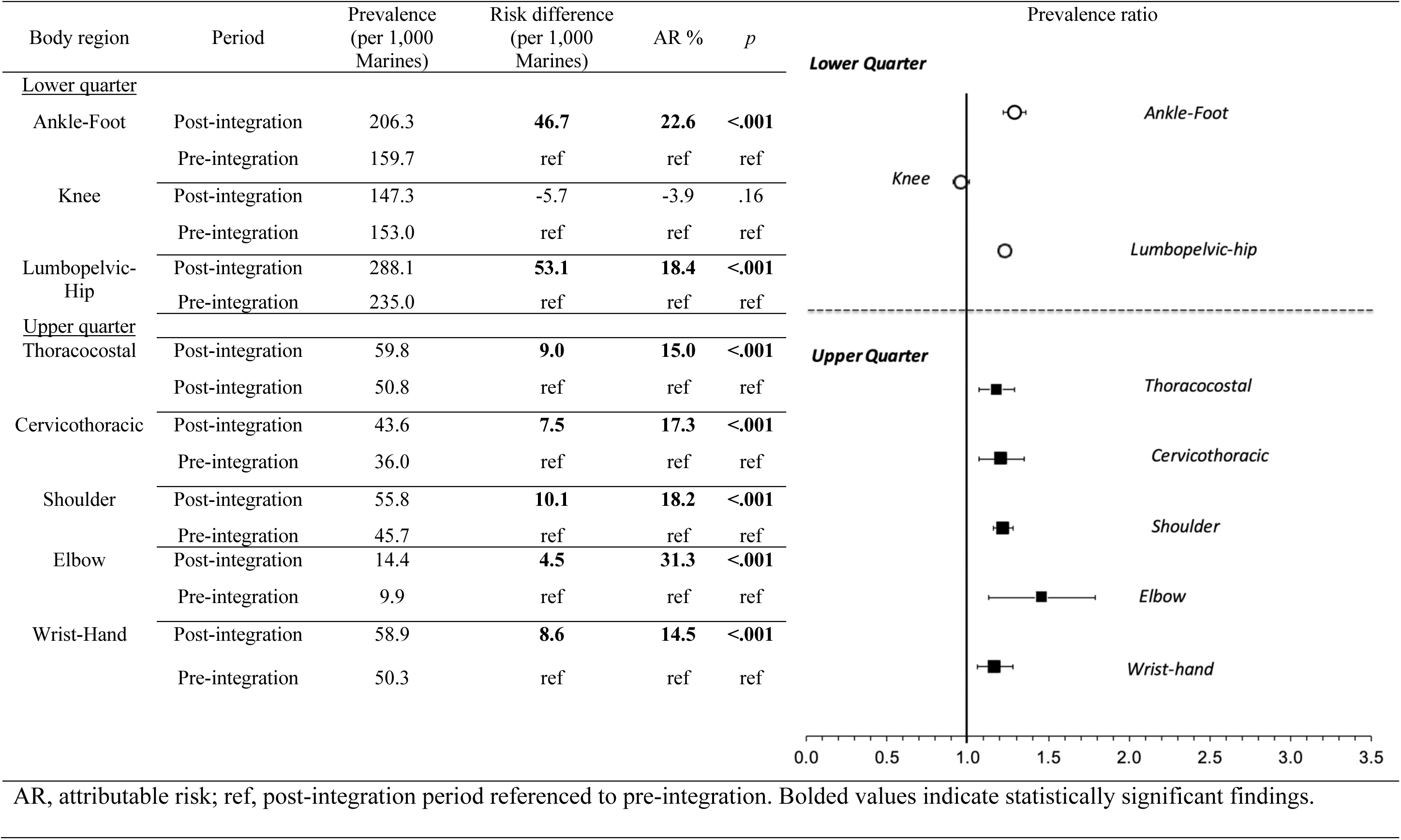
Assessment of Musculoskeletal Injury Risk in Female Marines, Stratified by Body Region, in the Post-Integration Period (2016–2020) Compared With the Pre-Integration Period (2011–2015)

Supplemental Figure 2 illustrates the direct acyclic graph detailing the effects of sex and occupation with moderation of integration period on musculoskeletal injury. In the multivariable evaluation of sex, occupation, integration period, and the interaction of sex and occupation on combined MSKI, only female sex was a salient factor for injury (PR = 1.99), with service in ground combat and aviation occupations identified as protective factors compared with services occupations (PR = 0.69) (Table 4). When these same factors were evaluated in the risk for specific MSKIs, female sex remained a robust and salient factor in all lower quarter injuries (PR = 1.75–2.63) and upper quarter injuries (PR = 1.38–2.36) except for shoulder injuries (Supplemental Tables 1 and 2). Service in ground combat and aviation occupations were protective for all lower quarter injuries (PR = 0.46–0.71). In the upper quarter, ground combat was protective for all but elbow injuries (PR = 0.67–0.77). Serving as an aviator was a risk factor for cervicothoracic (PR = 1.57) and thoracocostal (PR = 1.22) injuries and a protective factor for shoulder (PR = 0.73) and wrist-hand (PR = 0.46) injuries. Less than half of the upper and lower quarter MSKI categories were significantly different when comparing ground combat support and aviation support with services occupations. Adjusted risk for lumbopelvic-hip (PR = 1.13), ankle-foot (PR = 1.53), cervicothoracic (PR = 1.19), thoracocostal (PR = 1.14), and elbow (PR = 1.48) injuries significantly increased during the post-integration period. There was a significant sex-by-period interaction for shoulder injuries alone, with female sex in the post-integration epoch found to be salient (PR = 1.26).

**Table 4.** Results of the Multivariable Negative Binomial Regression Assessing Sex, Occupation, Integration Period, and Sex-by-Period Interaction on Combined Musculoskeletal Injury Risk, 2011–2020

|  |  | RR | 95% CI |  | *p* |
| --- | --- | --- | --- | --- | --- |
|  |  |  | LL | UL |  |
| Sex | Female | **1.99** | **1.78** | **2.23** | **<.001** |
| Occupation | Ground combat | **0.69** | **0.58** | **0.84** | **<.001** |
|  | Ground combat support | 0.94 | 0.80 | 1.11 | .48 |
|  | Aviation | **0.69** | **0.57** | **0.82** | **<.001** |
|  | Aviation support | 0.95 | 0.80 | 1.12 | .54 |
| Period | Post-integration (2016–2020) | **1.21** | **1.08** | **1.35** | **<.001** |
| Sex-by-period interaction | Female sex, post-integration | Non-significant |  |  |  |
| Negative log-likelihood ratio |  |  | 5449 |  |  |
| Reference groups: sex (male); occupation (services); period (pre-integration, 2011–2016); sex-by-occupation interaction (male Marines in services occupations); sex-by-period interaction (male Marines during the pre-integration period). RR, relative risk; LL, lower limit; UL, upper limit. Bolded values indicate statistically significant findings. |  |  |  |  |  |

## DISCUSSION

The primary findings of this study were that female sex was a salient factor in both the pre-integration and post-integration periods for overall MSKI risk. During the post-integration period, female sex was a salient factor for all lower quarter injuries and thoracocostal, cervicothoracic, and wrist-hand injuries compared with male Marines. There was a higher prevalence of ankle-foot and lumbopelvic-hip injuries in the lower quarter and all upper quarter injuries in women following integration. Once adjusted for occupation, integration period, and the interaction of sex and occupation on overall MSKI risk, female sex was a salient factor for injury, with service in ground combat and aviation occupations identified as protective factors compared with services occupations. In the evaluation of specific MSKIs, female sex remained a robust and salient factor in all lower quarter injuries and upper quarter injuries except for shoulder injuries. There was only a significant sex-by-period interaction for shoulder conditions, with an increased risk of these injuries in female Marines in the post-integration period. This is the first study, to our knowledge, to have evaluated MSKI risk in female service members serving in the Fleet Marine Force following integration.

Our finding that female Marines have a significant higher overall risk of MSKI compared with their male counterparts is consistent with previous studies.^1,20^ The prevalence of MSKI conditions significantly increased during the post-integration period in all body regions except for knee injuries. In the post-integration period, there was a noted reduction in injuries during 2020, a finding consistent with another study reporting the effects of the pandemic on MSKI risk.^1^ When adjusted MSKI risk was assessed with consideration of military occupation, female sex continued to be a robust and significant factor. Despite increased prevalence of MSKI in both female and male Marines during the post-integration period, the interaction of sex and period was not significant.

Among the occupations, the combat and aviator occupational communities had a significantly lower risk of MSKI once sex was factored in. This was consistent among all lower extremity, thoracocostal, lumbopelvic-hip, and shoulder injuries. The risk of MSKI in the combat support and aviation support occupations was not significantly different from that observed in services. It is plausible that there are cultural or geographic factors for increased diagnosis of these conditions in the support and services occupations. It is plausible that support and service fields are closer to medical support (fewer geographic barriers to care) and have occupational cultures that are more accepting and facilitate medical care-seeking.^12,13^ Ground and aviation combat occupations attract Marines that are resilient and more likely to self-manage symptoms. By avoiding care-seeking, perceived threat of being removed from operational duties (e.g., patrol or grounding from flight) and the appearance of weakness can be avoided.^12,13^ These suppositions, including care-seeking behaviors for MSKIs among female and male Marines serving in the different occupational communities, warrant further investigation.

### Clinical and Policy Implications

The findings in the current study can be used to plan mitigation strategies needed to ameliorate risk factors and the specialized medical capabilities needed to address these conditions in the Marines Corps. This is especially salient as more women have increasing involvement in ground combat operations. These findings further illustrate the need for surveillance and prevention programs “left of boom,” and integrated musculoskeletal specialists, including sports medicine physicians, physical therapists, and athletic trainers for in-garrison care. Once deployed, increased numbers of uniformed Navy sports medicine physicians and physical therapists are needed to provide prompt primary care to Marines in the austere operational environment while shipboard and when projected ashore. This recommendation is a stark departure from the current medical capability provided for Marines. While there are some specialized capabilities provided by Navy Medicine (Sports Medicine and Reconditioning Team; Navy Physical Therapy MSKI) and the Marine Corps (Marine Corps Sports Medicine Injury Prevention) for Marines in garrison, expansion of the interdisciplinary sports medicine programs is needed. Furthermore, there are no established Marine Corps requirements or doctrine that provide for non-trauma MSKI care in Role I–III settings. Establishment of specific requirements, integration into the medical annex of operational plans, and proper resourcing (i.e., Navy Sports Medicine Physicians and Physical Therapists in the table of organization) will ensure both female and male Marines are ready to fight while in garrison, afloat, and when projected ashore in the operational environment.

### Limitations

There are limitations to this study. First, estimates of MSKIs relied on healthcare encounters in the Medical Data Repository. Since undocumented care, documented care that was privately procured external to the healthcare benefit, or self-care would not be reflected in our results, the true prevalence of MSKI in the Marine Corps is likely much greater than that reported in this study. This is especially true in the ground and aviation combat occupations, where it is estimated that approximately 37% of Marines do not seek care for these injuries.^12^ The epoch employed during the study occurred during a period of relative peacetime. It is unclear if these findings would be influenced by large-scale conflict, which would limit generalizability.

## CONCLUSION

Female sex was a salient factor for MSKI, with service in ground combat and aviation occupations identified as protective factors when compared with services occupations. In the evaluation of specific MSKIs, female sex remained a robust and salient factor in all lower quarter injuries and upper quarter injuries except for shoulder injuries. There was only a significant sex-by-period interaction for shoulder conditions, with an increased risk of these injuries in female Marines in the post-integration period.

## Declarations

Dr. Fraser reports grants from Congressionally Directed Medical Research Programs and the Office of Naval Research, outside of the submitted work. In addition, Dr. Fraser has a patent pending for an Adaptive and Variable Stiffness Ankle Brace, U.S. Provisional Patent Application No. 63254,474.

## Acknowledgments

None

## Prior Presentation

This work was presented at the 2024 Military Health System Research Symposium, August 2024, Kissimmee, Florida.

## Clinical Trial Registration

Not applicable

## Institutional Review Board (Human Subjects)

The study protocol was approved by the Naval Health Research Center Institutional Review Board in compliance with all applicable federal regulations governing the protection of human subjects. Research data were derived from approved Naval Health Research Center Institutional Review Board protocol number NHRC.2003.0025.

## Institutional Animal Care and Use Committee (IACUC)

Not applicable

## Individual Author Contributions

JJF and MCH were responsible for study conception and methodological development. JJF and JMZ contributed to data analysis. All authors contributed to the data interpretation, manuscript development, critical revision, and final approval of the study.

## Data Availability

The datasets generated and/or analyzed during the current study are not publicly available due to personally identifiable information regulations, but they may be made available by the corresponding author on reasonable request and approval by the Naval Health Research Center Institutional Review Board/Privacy Office.

## Disclaimer

JJF, JMZ, PHS, and AJM are employees of the U.S. Government and this work was prepared as part of their official duties. Title 17, U.S.C. §105 provides that copyright protection under this title is not available for any work of the U.S. Government. Title 17, U.S.C. §101 defines a U.S. Government work as work prepared by a military service member or employee of the U.S. Government as part of that person’s official duties. Report No. 25-55 was supported by the Office of Naval Research (grant number N00014-21-12892) under work unit no. 60808. The views expressed are those of the authors and do not necessarily reflect the official policy or position of the Uniformed Services University of the Health Sciences, the Department of the Navy, Department of Defense, the Department of Veterans Affairs, nor the U.S. Government.

## Institutional Clearance

Institutional clearance approved.

## Supplemental Material

**Supplemental Figure 1.**
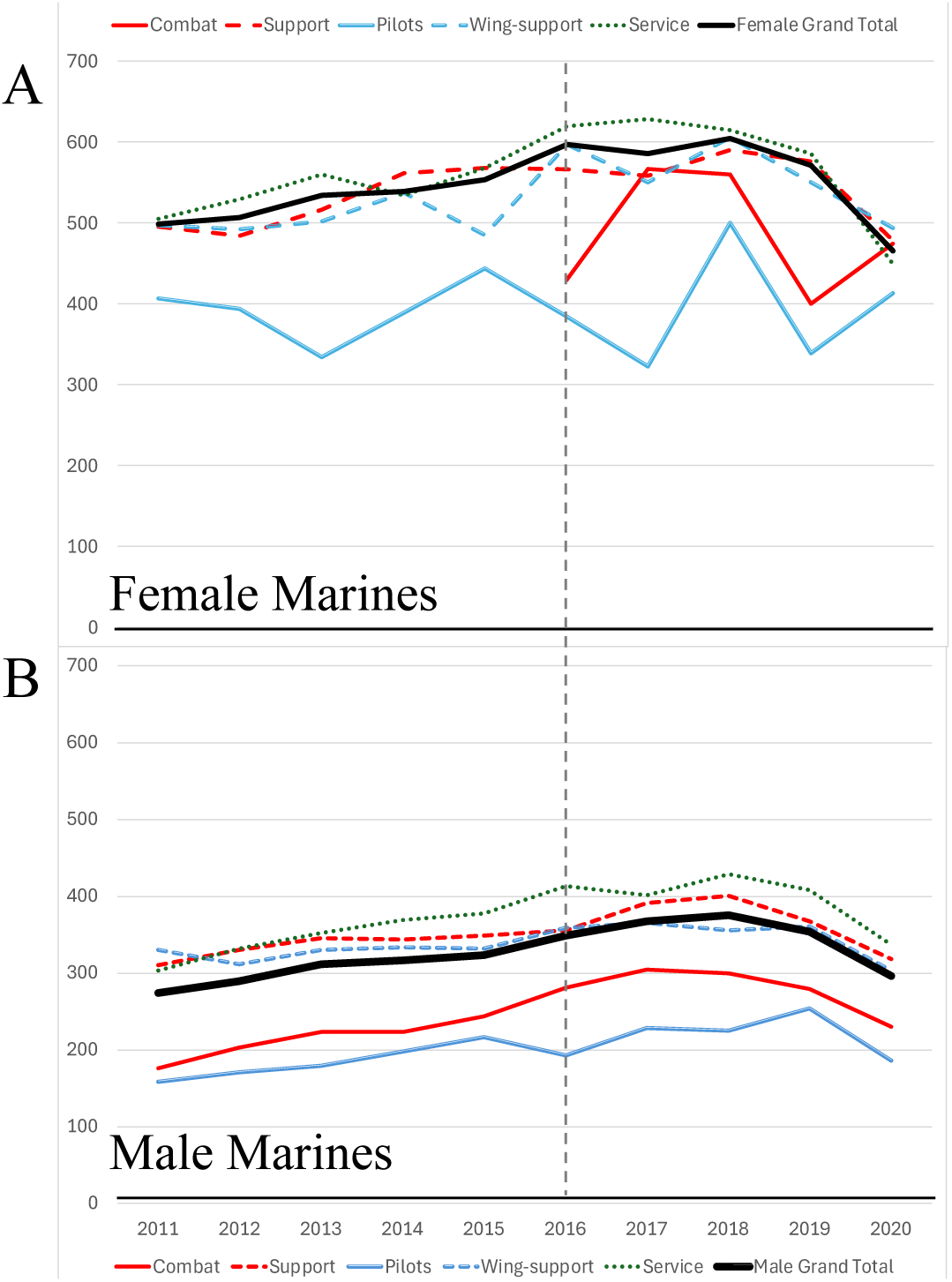
Prevalence of musculoskeletal injuries over time in male and female Marines serving in each occupational category and for all occupations combined.

**Supplemental Figure 2.**
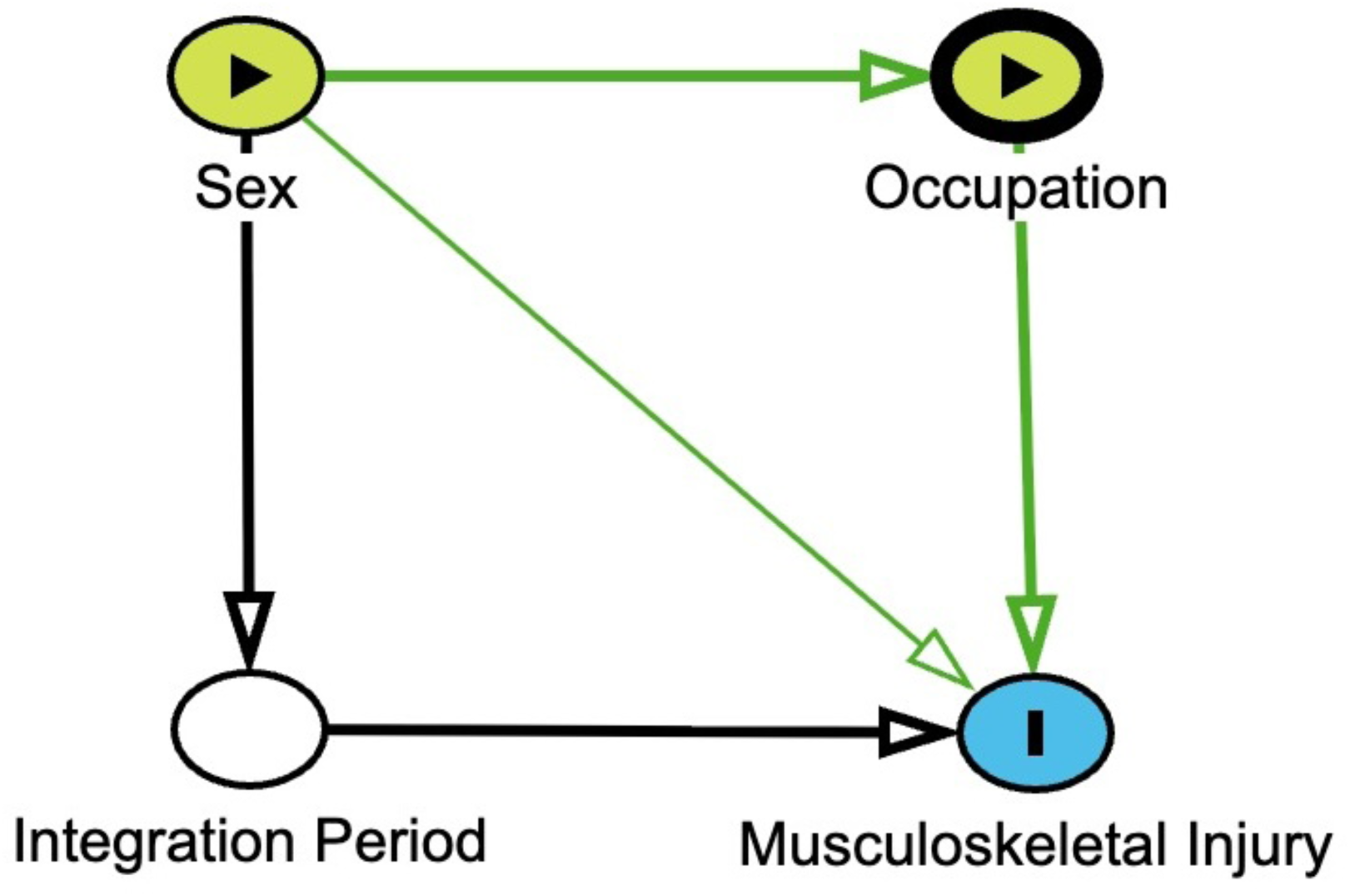
Direct acyclic graph detailing the effects of sex and occupation with moderation of integration period on musculoskeletal injury.

**Supplemental Table 1.** Results of the multivariable negative binomial regression assessing sex, occupation, integration period, and sex by period interaction on lower quarter injury risk in US Marines, 2011-2020.

|  |  | <u>Lumbopelvic-Hip</u> |  |  |  | <u>Knee</u> |  |  |  | <u>Ankle-Foot</u> |  |  |  |
| --- | --- | --- | --- | --- | --- | --- | --- | --- | --- | --- | --- | --- | --- |
|  |  | RR | 95% CI |  | *p* | RR | 95% CI |  | *p* | RR | 95% CI |  | *p* |
|  |  |  | LL | UL |  |  | LL | UL |  |  | LL | UL |  |
| Sex | Female | **2.63** | **2.38** | **2.90** | **<.001** | **1.75** | **1.57** | **1.95** | **<.001** | **1.92** | **1.72** | **2.14** | **<.001** |
| Occupation | Ground combat | **0.71** | **0.60** | **0.83** | **<.001** | **0.61** | **0.51** | **0.73** | **<.001** | **0.64** | **0.53** | **0.76** | **<.001** |
|  | Ground combat support | 0.98 | 0.85 | 1.13 | .79 | 0.96 | 0.81 | 1.12 | .58 | 0.90 | 0.77 | 1.07 | .23 |
|  | Aviation | **0.63** | **0.54** | **0.73** | **<.001** | **0.46** | **0.39** | **0.54** | **<.001** | **0.53** | **0.45** | **0.63** | **<.001** |
|  | Aviation support | 1.15 | 0.99 | 1.33 | .07 | **0.85** | **0.72** | **0.99** | **.04** | **0.78** | **0.66** | **0.92** | **.003** |
| Period | Post-integration (2016–2020) | **1.13** | **1.02** | **1.25** | **.02** | 1.05 | 0.94 | 1.17 | .37 | **1.53** | **1.37** | **1.71** | **<.001** |
| Sex-by-period interaction |  | Non-significant |  |  |  | Non-significant |  |  |  | Non-significant |  |  |  |
| Negative log-likelihood ratio |  | 672.5 |  |  |  | 650.2 |  |  |  | 668.7 |  |  |  |

Reference groups: sex (male); occupation (services); period (pre-integration, 2011–2016); sex-by-occupation interaction (male Marines in services occupations); sex-by-period interaction (male Marines during the pre-integration period). RR, relative risk; LL, lower limit; UL, upper limit. Bolded values indicate statistically significant findings.

**Supplemental Table 2.**
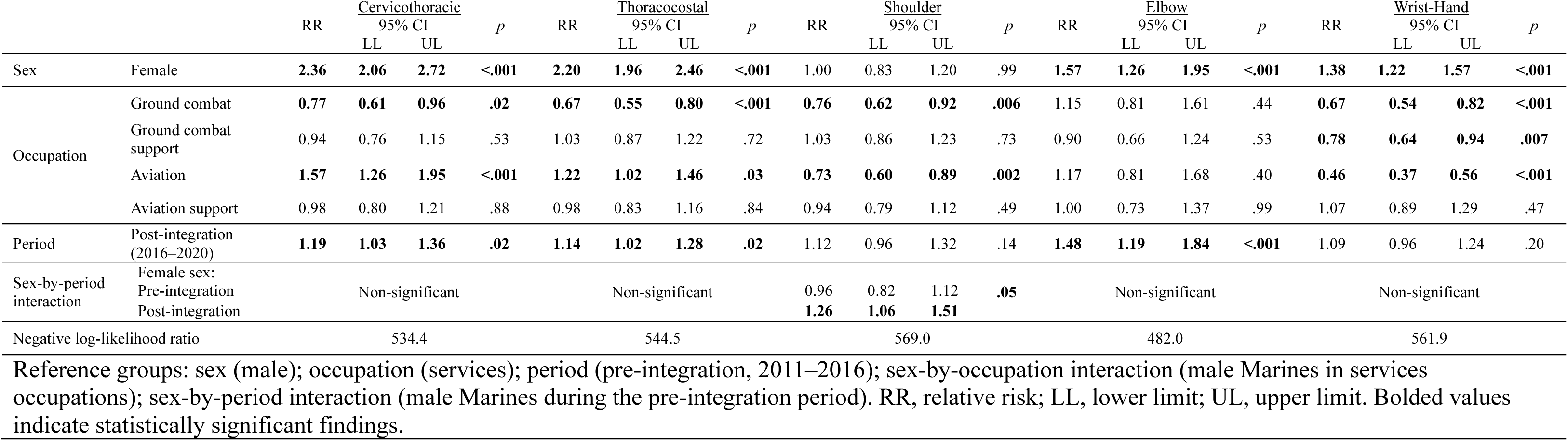
Results of the Multivariable Negative Binomial Regression Assessing Sex, Occupation, Integration Period, and Sex-by-Interaction on Lower Quarter Injury Risk in U.S. Marines, 2011–2020

